# Are the SIR and SEIR models suitable to estimate the basic reproduction number for the CoViD-19 epidemic?

**DOI:** 10.1101/2020.10.11.20210831

**Authors:** Hyun Mo Yang, Luis Pedro Lombardi Junior, Ariana Campos Yang

## Abstract

The transmission of severe acute respiratory syndrome coronavirus 2 (SARS-CoV-2) becomes pandemic but presents different incidences in the world. Mathematical models were formulated to describe the coronavirus disease 2019 (CoViD-19) epidemic in each country or region. At the beginning of the pandemic, many authors used the SIR (susceptible, infectious, and recovered compartments) and SEIR (including exposed compartment) models to estimate the basic reproduction number *R*_0_ for the CoViD-19 epidemic. These simple deterministic models assumed that the only available collection of the severe CoViD-19 cases transmitted the SARS-CoV-2 and estimated lower values for *R*_0_, ranging from 1.5 to 3.0. However, the major flaw in the estimation of *R*_0_ provided by the SIR and SEIR models was that the severe CoViD-19 patients were hospitalized, and, consequently, not transmitting. Hence, we proposed a more elaborate model considering the natural history of CoViD-19: the inclusion of asymptomatic, pre-symptomatic, mild and severe CoViD-19 compartments. The model also encompassed the fatality rate depending on age. This SEAPMDR model estimated *R*_0_ using the severe CoViD-19 data from São Paulo State (Brazil) and Spain, yielding higher values for *R*_0_, that is, 6.54 and 5.88, respectively. It is worth stressing that this model assumed that severe CoViD-19 cases were not participating in the SARS-CoV-2 transmission chain. Therefore, the SIR and SEIR models are not suitable to estimate *R*_0_ at the beginning of the epidemic by considering the isolated severe CoViD-19 data as transmitters.

## 1 Introduction

The first case of coronavirus disease 2019 (CoViD-19) was detected in China in December 2019 and spread rapidly to other countries. In March 2020, the World Health Organization (WHO) declared the CoViD-19 a pandemic. The CoViD-19 is caused by the severe acute respiratory syndrome coronavirus 2 (SARS-CoV-2), which can be transmitted by droplets that escape the lungs by coughing or sneezing and infects humans through the nose, mouth, or eyes. In severe cases, immune cells overreact, causing acute respiratory disease syndrome and possibly death.

At the beginning of the epidemic, we have two data sets: Severe CoViD-19 cases (those in hospitals were tested and confirmed) and deaths. Due to the lack of mass testing (PCR and serology), the epidemic curve consisted of severe CoViD-19 cases. Mathematical models, usually the SEIR (susceptible, exposed, infectious and recovered compartments) and the simplified SIR, were used to describe this epidemic curve. The model parameters were fitted to estimate the basic reproduction number *R*_0_.

In the literature, the usually estimated or assumed basic reproduction number *R*_0_ is around 2.0. For instance, Koo *et al*. [1] and Ferguson *et al*. [2] assumed *R*_0_ ranging between 1.5 and 2.6 and simulated agent-based models to predict the number of CoViD-19 cases and deaths. However, Li *et al*. [3], based on the SEIR-metapopulations model, estimated the effective reproduction number *R*_*ef*_ considering data from January 10 to February 8, arguing that the most recent common ancestor could have occurred on November 17, 2019. The time elapsed from November 17, 2019 (the first case) to January 10, 2020 (the first day in the estimation) is 54 days. Moreover, on January 23, 2020, Wuhan and other Hubei province cities imposed a rigid lockdown. Hence, their estimation of *R*_*ef*_ = 2.38 considered the range of data recorded from 54 days after the epidemic’s onset to 16 days after the lockdown. The definition of the basic reproduction number is “one infectious individual is introduced in a completely susceptible population without constraints (interventions)” [4]. Therefore, the estimation of *R*_0_ must consider the severe CoViD-19 data restricted on the period without interventions.

The estimation of the basic reproduction number *R*_0_ depends on the mathematical model [5]. To exemplify this dependency to describe the CoViD-19 epidemic, we consider the SIR and SEIR models, plus an additional model taking into account more compartments and encompassing the fatality rate depending on the age. The latter model divides a population into two subpopulations and split the infectious (*I*) compartment into asymptomatic (*A*), pre-diseased or pre-symptomatic (*P*), mild (*M*), and severe (*D*) CoViD-19 compartments. This model [6], called the SEAPMDR model, was proposed to show fewer deaths due to severe CoViD-19 cases than those provided by the SIR and SEIR models [2]. However, the SEAPMDR model estimated higher values for *R*_0_.

The reliable estimation of *R*_0_ is essential because this number determines the magnitude of effort to eradicate infection. For instance, a vaccination must immunize a fraction equal to or greater than 1 − 1*/R*_0_ of the susceptible population to eradicate a disease [4]. In [7], analyzing vaccination as a control mechanism, if *R*_*ef*_ is reduced lower than one, the number of cases decreased following an exponential-type decay. Instead of vaccination, if we consider the isolation of susceptible individuals, for a lower value of *R*_0_ as provided by the SIR and SEIR models, a small fraction (1 − 1*/R*_0_) of the population must be in quarantine to control the transmission of SRS-CoV-2. However, the SEAPMDR model considering more details of the natural history of CoViD-19 estimated a higher value to *R*_0_, showing that the control is not an easy task.

As we have pointed out, only severe CoViD-19 cases were registered at the beginning of the epidemic. In the SIR and SEIR models, the collection of severe CoViD-19 data were allocated in the transmitting compartment *I* to estimate model parameters. However, in the SEAPMDR model, the observed data are allocated in the non-transmitting compartment *D*. Considering the severe CoViD-19 data collection from São Paulo State (Brazil) [8] and Spain [9], we fit the parameters of those models to calculate *R*_0_. We conclude that SIR and SEIR models are not suitable to estimate the basic reproduction number. For instance, the lower estimated values for *R*_0_ are a result of considering the data collected later after the beginning of the epidemic and data collected after the intervention (quarantine).

The paper is structured as follows. In Section 2, we describe SIR, SEIR, and SEAPMDR models used to estimate *R*_0_ in Section 3. Discussion is presented in Sections 4, and the conclusion in Section 5.

## 2 Material and methods

We present the SIR, SEIR, and SEAPMDR models in this section. From these models, we obtained *R*_0_ from the steady-state analysis.

### 2.1 The SIR and SEIR model

In the SEIR model, the population is divided into susceptible (*S*), exposed (*E*), infectious (*I*), and recovered (*R*) subpopulations (compartments). The population’s vital dynamics is described by the per-capita birth *ϕ* and natural mortality *µ* rates. In severe CoViD-19 cases, the overreacted immune cells may result in death described by the additional mortality (fatality) rate *α*.

According to the CoViD-19 natural history, susceptible individuals are infected at a rate *λS*, where the force of infection *λ* is given by

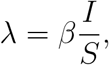

with *β* being the transmission rate. After a period *σ*^−1^, where *σ* is the incubation rate, these individuals enter into infectious compartment and, after a period *γ*^−1^, where *γ* is the infectious (or recovery) rate, they enter into recovered compartment. Hence, the SEIR model is described by the dynamic system

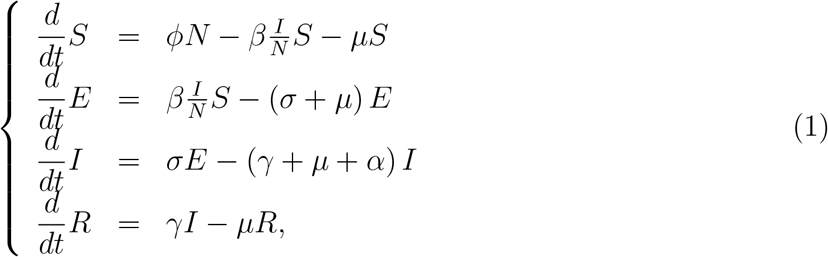

where the total population *N* = *S* + *E* + *I* + *R* obeys

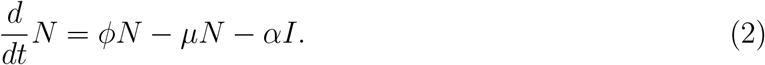

The number of accumulated severe CoViD-19 cases Ω is given by the exit from *E* and entering into class *I*, that is,

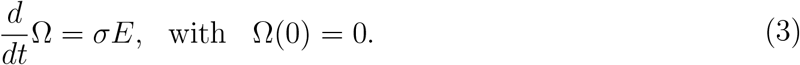

and this equation is accopled to the system of equations (1) to obtain numerically Ω.

The system of equations (1) is autonomous but describes non-constant population dynamics (if *ϕ* = *µ* + *αI/N*, then *N* is constant). This varying population is the reason why the equation (1) can not be used directly to obtain the basic reproduction number. In Appendix A.1, the steady-state of the system of equations in terms of the fractions corresponding to equation (1) was analyzed to obtain the basic reproduction number *R*_0_.

The SIR model is obtained letting *σ* → ∞, resulting in

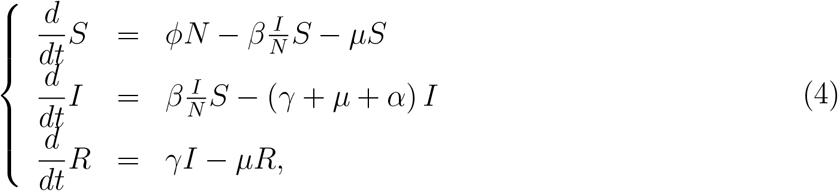

and the number of accumulated severe CoViD-19 cases Ω is given by the exit from *S* and entering into class *I*, that is,

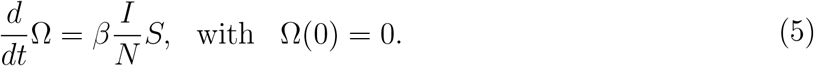

In Appendix A.2, the steady-state of the system of equations in terms of the fractions corresponding to equation (4) was analyzed to obtain the basic reproduction number *R*_0_.

## 2.2 The SEAPMDR model

One of the main aspects of CoViD-19 is increased fatality in the elder subpopulation. For this reason, a population is divided into two groups, composed of young (60 years old or less, denoted by subscript *y*) and elder (60 years old or more, denoted by subscript *o*) subpopulations. This community’s vital dynamic is described by the per-capita rates of birth (*ϕ*) and mortality (*µ*), and *ϕ* is the aging rate, that is, the flow from young subpopulation *y* to elder subpopulation *o*. Another aspect is the presence of the pre-symptomatic individuals, that is, individuals without symptoms transmitting SARS-CoV-2 before the onset of the disease [10].

Since we are dealing with the initial phase of the epidemic, the model does not consider the compartments related to quarantine and mass testing. Hence, for each subpopulation *j* (*j* = *y, o*), individuals are divided into six classes: susceptible *S*_*j*_, exposed and incubating *E*_*j*_, asymptomatic *A*_*j*_, pre-symptomatic (or pre-diseased) individuals *P*_*j*_, symptomatic individuals with mild CoViD-19 *M*_*j*_, and severe CoViD-19 *D*_*j*_. However, all young and elder individuals in classes *A*_*j*_, *M*_*j*_, and *D*_*j*_ enter into the same recovered class *R* (this is the 7^*th*^ class, but common to both subpopulations). Hence, the SEAPMDR model has 13 compartments.

The natural history of CoViD-19 is the same for young (*j* = *y*) and elder (*j* = *o*) subpopulations. We assume that individuals in the asymptomatic (*A*_*j*_), pre-diseased (*P*_*j*_), and a fraction *z*_*j*_ of mild CoViD-19 (*M*_*j*_) classes are transmitting the virus. Other infected classes ((1 − *z*_*j*_) *M*_*j*_ and *D*_*j*_) are under voluntary or forced isolation. Susceptible individuals are infected at a rate *λ*_*j*_*S*_*j*_ (known as the mass action law [4]), where *λ*_*j*_ is the per-capita incidence rate (or force of infection) defined by *λ*_*j*_ = *λ* (*δ*_*jy*_ + *ψδ*_*jo*_), with *λ* being

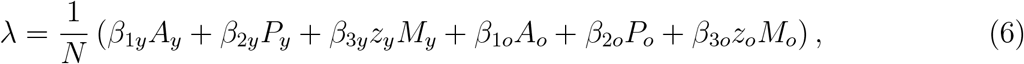

where *δ*_*ij*_ is the Kronecker delta, with *δ*_*ij*_ = 1 if *i* = *j*, and 0, if *i* ≠ *j*; and *β*_1*j*_, *β*_2*j*_ and *β*_3*j*_ are the transmission rates, that is, the rates at which a virus encounters a susceptible people and infects him/her. In [6], a particular model was analyzed letting *z*_*y*_ = *z*_*o*_ = 0 and *χ*_*y*_ = *χ*_*o*_ = 1.

Susceptible individuals are infected at a rate *λ*_*j*_ and enter into class *E*_*j*_. After an average period 1*/σ*_*j*_ in class *E*_*j*_, where *σ*_*j*_ is the incubation rate, exposed individuals enter into the asymptomatic class *A*_*j*_ (with probability *l*_*j*_) or pre-diseased class *P*_*j*_ (with probability 1 − *l*_*j*_). After an average period 1*/γ*_*j*_ in class *A*_*j*_, where *γ*_*j*_ is the recovery rate of asymptomatic individuals, asymptomatic individuals acquire immunity (recovered) and enter into recovered class *R*. Possibly asymptomatic individuals can manifest symptoms at the end of this period, and a fraction 1 − *χ*_*j*_ enters into mild CoViD-19 class *M*_*j*_. For symptomatic individuals, after an average period 1*/γ*_1*j*_ in class *P*_*j*_, where *γ*_1*j*_ is the infection rate of pre-diseased individuals, pre-diseased individuals enter into severe CoViD-19 class *D*_*j*_ (with probability 1 − *k*_*j*_) or mild CoViD-19 class *M*_*j*_ (with probability *k*_*j*_). Individuals in class *D*_*j*_ acquire immunity after a period 1*/γ*_2*j*_, where *γ*_2*j*_ is the recovery rate of severe CoViD-19, and enter into recovered class *R* or die under the disease-induced (additional) mortality rate *α*_*j*_. Individuals in mild CoViD-19 class *M*_*j*_ acquire immunity after a period 1*/γ*_3*j*_, where *γ*_3*j*_ is the recovery rate of mild CoViD-19, and enter into recovered class *R*.

The SARS-CoV-2 transmission model is described by the system of ordinary differential equations. Equations for susceptible individuals are

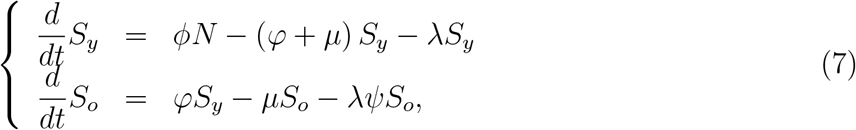

for infectious individuals, with *j* = *y, o*,

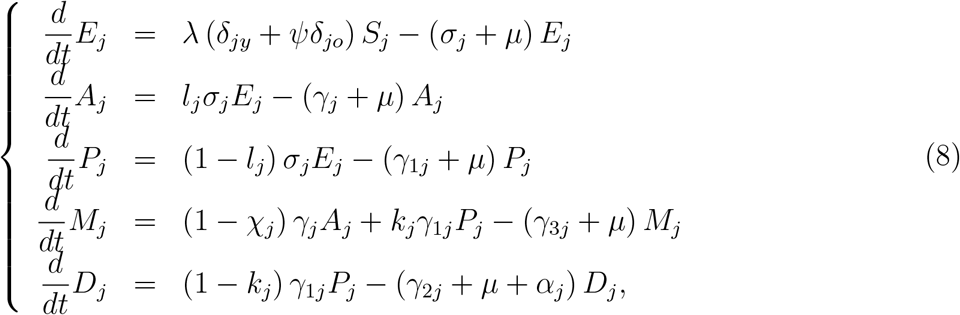

and for recovered individuals,

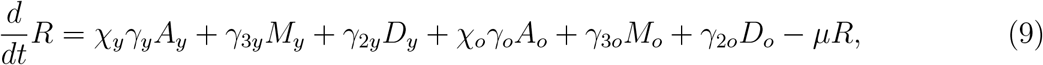

where *N*_*j*_ = *S*_*j*_ + *E*_*j*_ + *A*_*j*_ + *P*_*j*_ + *M*_*j*_ + *D*_*j*_, and *N* = *N*_*y*_ + *N*_*o*_ + *I* obeys

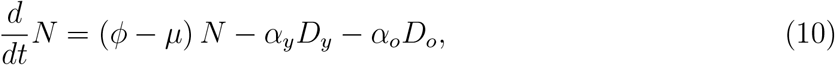

with the initial number of population at *t* = 0 being *N* (0) = *N*_0_ = *N*_0*y*_ + *N*_0*o*_, where *N*_0*y*_ and *N*_0*o*_ are the size of young and elder subpopulations at *t* = 0.

The number of accumulated severe CoViD-19 cases Ω is obtained from

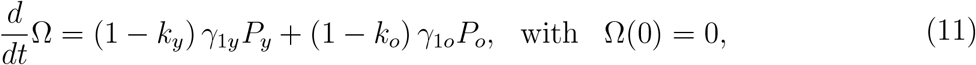

which are the exit from class *P*, and entering into class *D*.

Table 1 summarizes the model parameters. The description of the assigned values can be found in [11]. The transmission rates are estimated.

**Table 1:**
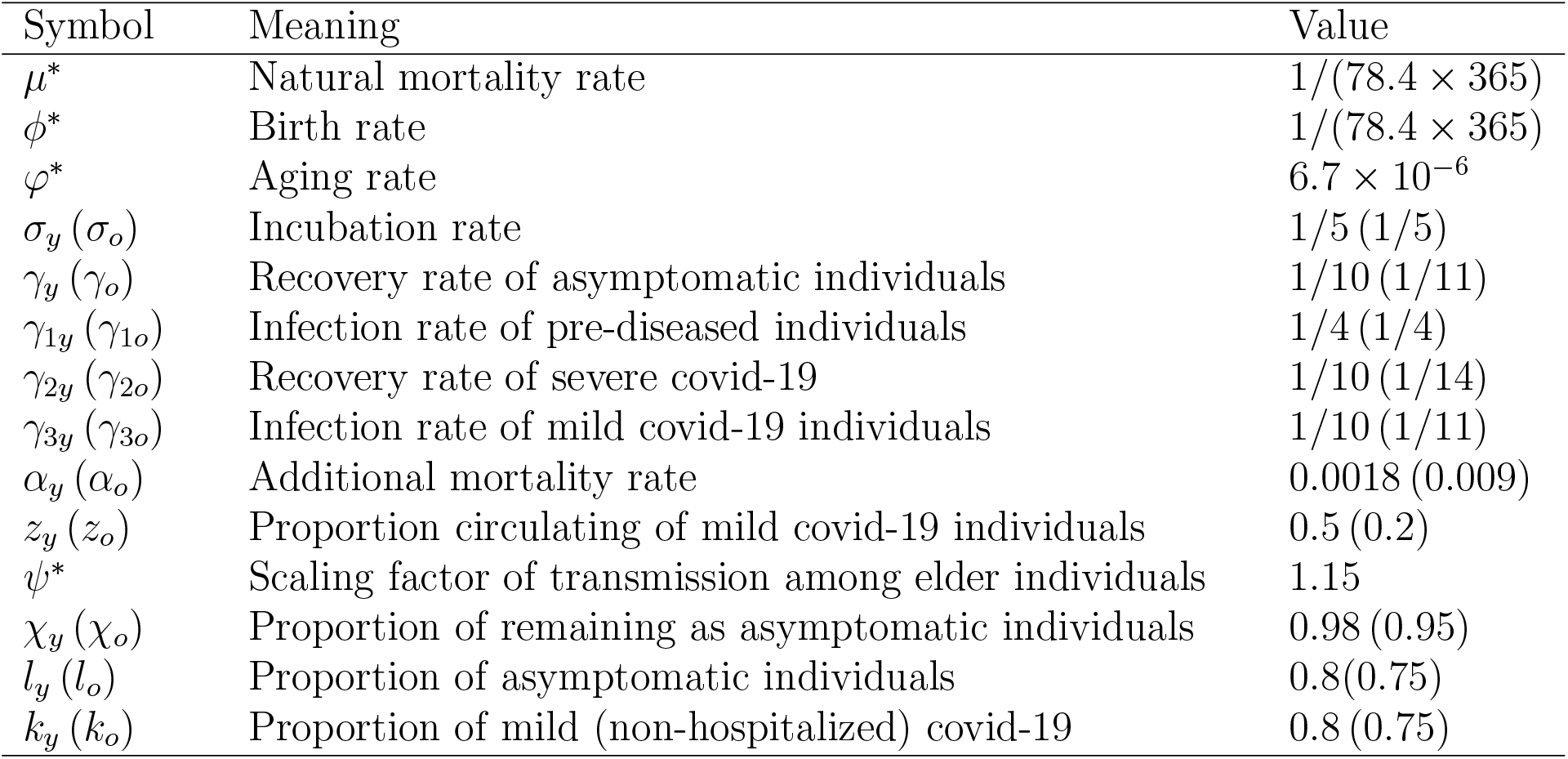
Summary of the model parameters (*j* = *y, o*) and values (rates in *days*^−1^, and proportions are dimensionless). The values (^*∗*^) correspond to São Paulo State. For Spain, *ϕ* = *µ* = 1*/*(83.4 × 365) *days*^−1^, *φ* = 1.14 × 10^−5^ *days*^−1^, and *ψ* = 1.1.

In Appendix B, the steady-state of the system of equations in terms of fractions corresponding to equations (7), (8) and (9) was analyzed to obtain the basic reproduction number *R*_0_.

The basic reproduction number *R*_0_ given by equation (B.8) in Appendix B, with the fractions written as 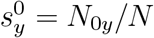 and 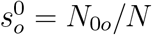, is

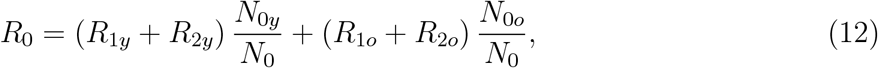

where *N*_0*y*_ and *N*_0*o*_ are the initial numbers of young and elder subpopulations with *N*_0_ = *N*_0*y*_ + *N*_0*o*_, and

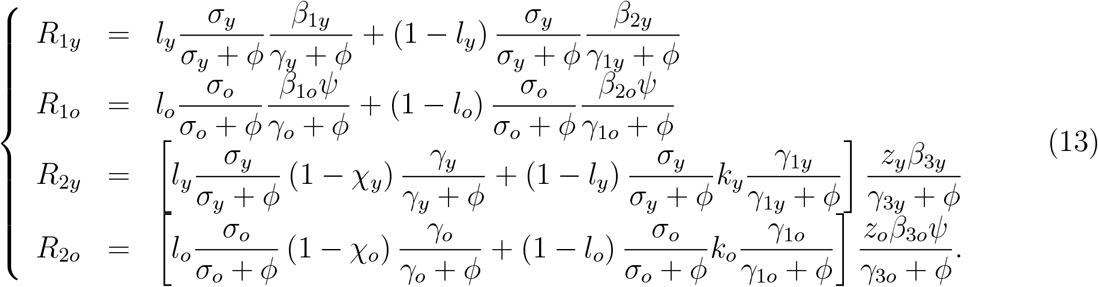

Letting *z*_*y*_ = *z*_*o*_ = 0 (*R*_2*y*_ = *R*_2*o*_ = 0), we retrieve the basic reproduction number obtained in [11].

### 2.3 The sigmoid-shaped curve of Ω

The first derivative of Ω is given by equations (5), (3) and (11) for, respectively, SIR, SEIR and SEAPMDR models, or

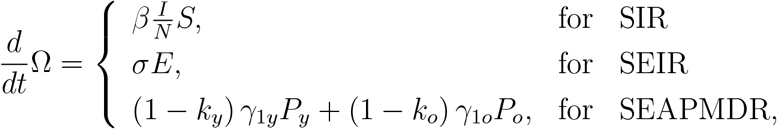

which is positive (*d*Ω*/dt* > 0). Hence, Ω is monotonically increasing function.

The second derivative of Ω is given

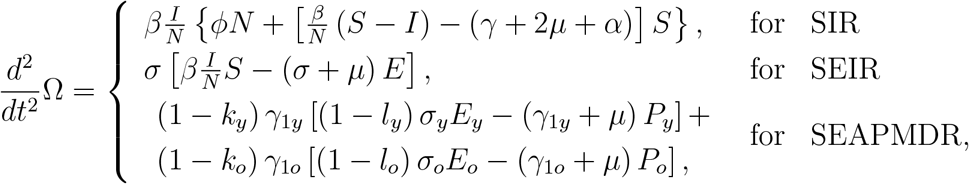

which changes the signal from positive to negative at *d*^2^Ω*/dt*^2^ = 0. Hence, Ω has upward concavity at the beginning of the epidemic and downward concavity at the ending phase of the epidemic. For instance, the number of exposed individuals *E* in the SEIR model initially increases (*dE/dt* > 0), attains a maximum value (*dE/dt* = 0), and decreases since after (*dE/dt* < 0). Hence, the change in the concavity occurs at the inflection time (point) *τ* satisfying *dE/dt* = 0, that is, *βIS/N* − (*σ* + *µ*) *E* = 0.

Therefore, the accumulated severe CoViD-19 cases Ω follows a sigmoid-shape [12]. From the first derivative of Ω given by equations (5), (3) and (11), we can retrieve the daily severe covid-19 cases Ω_*d*_ as

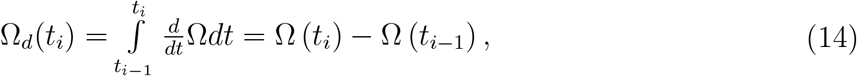

where Ω_*d*_(0) = 0 at *t*_0_ = 0, and Δ*t* = *t*_*i*_ − *t*_*i*−1_ = Δ*t* = 1 *day*, with *i* = 1, 2, …, and *t*_1_ = 1 is the next day in the calendar time, and so on. The inflection time *τ* occurs at the maximum value of Ω_*d*_.

The curve of the effective reproduction number *R*_*ef*_ ≈ *R*_0_*S/N* is given by

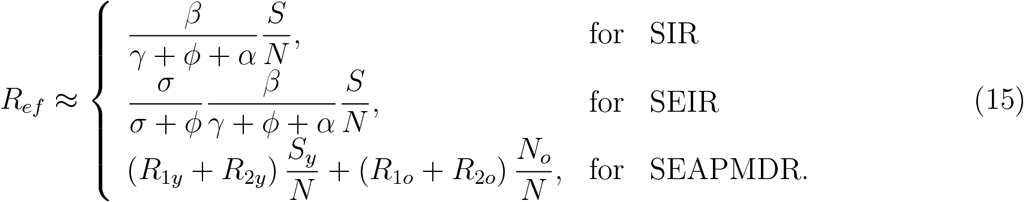

This curve can be used to explain the sigmoid-shape of the accumulated severe CoViD-19 cases Ω.

When *d*^2^Ω*/dt*^2^ > 0, the upward concavity curve of Ω increases quickly, which is possible if *R*_*ef*_ > 1, and one primary case generates more than one secondary case. However, when *d*^2^Ω*/dt*^2^ < 0, the downward concavity curve of Ω increases slowly reaching an asymptote, which is possible if *R*_*ef*_ < 1, and one primary case generates less than one secondary case. Hence, the effective reproduction number *R*_*ef*_ decreases monotonically from *R*_0_ at *t* = 0 up to one at the inflection time *τ* (upward concavity of Ω), and maintains its decreasing trend up to an asymptote (zero, if *µ* = 0), forming the phase of the downward concavity of the curve Ω. However, *R*_*ef*_ = 1 does not occur at the inflection time *τ* (see below).

## 3 Results

We estimate the transmission rate *β* for the SIR, SEIR, and SEAPMDR models presented in the preceding section against the severe CoViD-19 data collection. The basic reproduction number *R*_0_ is then calculated for São Paulo State (Brazil) and Spain.

São Paulo State has 44.6 million inhabitants (demographic density, 177*/km*^2^) with 15.3% of the population comprised of elder individuals [8]. The first case was registered on February 26, and partial quarantine was implemented on March 24. The sum of the incubation (1*/σ*) and pre-symptomatic (1*/γ*_1_) periods is 9 days, for this reason it is expected a delay of around 9 days since the infection and the CoViD-19 onset. Therefore, we estimate the basic reproduction number considering the collected data from February 26 to April 2.

Spain has 47.4 million inhabitants (demographic density, 92.3*/km*^2^) with 25.8% of the population comprised of elder individuals [9]. The first case was registered on January 31, and the lockdown was implemented on March 16. Considering a delay of around 9 days, we estimate the basic reproduction number considering the collected data from January 31 to March 25.

To estimate the parameter *β*, we calculate

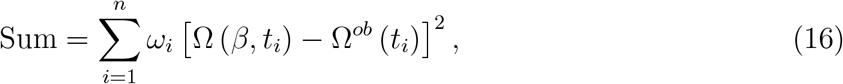

where Ω (*β, t*_*i*_) is the accumulated severe CoViD-19 cases calculated from the dynamic system (SIR, SEIR and SEAPMDR models), and Ω^*ob*^ (*t*_*i*_) is the accumulated severe CoViD-19 registered cases at day *t*_*i*_, that is,

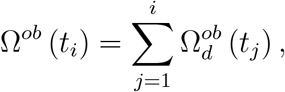

where 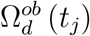 is the severe CoViD-19 cases registered at day *t*_*j*_. We consider the same weight for all data, that is, we let *ω*_*i*_ = 1. The value of *β* that minimizes Sum is the fitted value.

Instead of applying the least square estimation method, we calculate the sum of the squared differences between the calculated Ω and the observed data Ω^*ob*^. The reason behind this simplification relies on the fitting of the model parameter based on a unique observed variable (Ω) of the dynamic system, which is not appropriate [13]. However, this simplified method of parameter evaluation does not provide uncertainties associated with the parameters. The effects of these uncertainties on the epidemic can be assessed by the global sensitivity analysis [14].

Once the transmission rate *β* is estimated, we calculate *R*_0_ using equations (A.7), (A.3) and (B.8) for, respectively, SIR, SEIR and SEAPMDR models. Notice that these three expressions for *R*_0_ give secondary cases produced by one primary case introduced in a completely susceptible population. Notice that the fatality rate *α* affects *R*_0_ in the SIR and SEIR models, but not in the SEAPMDR model. The reason behind it is the severe CoViD-19 cases transmitting in the SIR and SEIR models but not in the SEAPMDR model. Hence, in the latter model, one of the sources of uncertainties is removed.

### 3.1 The SIR model

To estimate the transmission rate, we use equation (16) and Ω given by equation (5). The model parameters are *γ* = 1*/*10, *α* = 0.002 and, for São Paulo State, *ϕ* = *µ* = 1*/*(78.4 × 365) and, for Spain, *ϕ* = *µ* = 1*/*(83.4 × 365) (all in *days*^−1^). The value *γ* = 1*/*10 *days*^−1^ in somehow is an average value among *γ, γ*_1_, *γ*_2_, and *γ*_3_ in the SEAPMDR model (see Table 1). The transmission rate *β* is estimated, and the basic reproduction number *R*_0_ is calculated using equation (A.7). We estimate the basic reproduction number using equation (4) with different infective individuals at *t* = 0.

For the data collected from São Paulo State, we obtained for *I*(0) = 1, *R*_0_ = 3.14 with Sum = 8.02 × 10^5^, for *I*(0) = 10, *R*_0_ = 2.4 with Sum = 8.49 × 10^5^, for *I*(0) = 25, *R*_0_ = 2.11 with Sum = 1.86 × 10^6^, and for *I*(0) = 100, *R*_0_ = 1.62 with Sum = 5.87 × 10^6^. Other initial conditions are *S*(0) = 44.6 million and *R*(0) = 0. The lowest Sum occurs when *R*_0_ = 3.14.

For the data collected from Spain, we obtained for *I*(0) = 1, *R*_0_ = 2.97 with Sum = 4.16 × 10^8^, for *I*(0) = 10, *R*_0_ = 2.5 with Sum = 6.32 × 10^8^, for *I*(0) = 25, *R*_0_ = 2.3 with Sum = 1.13 × 10^9^, and for *I*(0) = 100, *R*_0_ = 2.06 with Sum = 1.19 × 10^9^. Other initial conditions are *S*(0) = 47.4 million and *R*(0) = 0. The lowest Sum occurs when *R*_0_ = 2.97.

Figure 1 shows the estimated curve Ω for São Paulo (a) and Spain (b) with three different initial conditions (*I*(0) = 1, 10, and 25).

**Figure 1:**
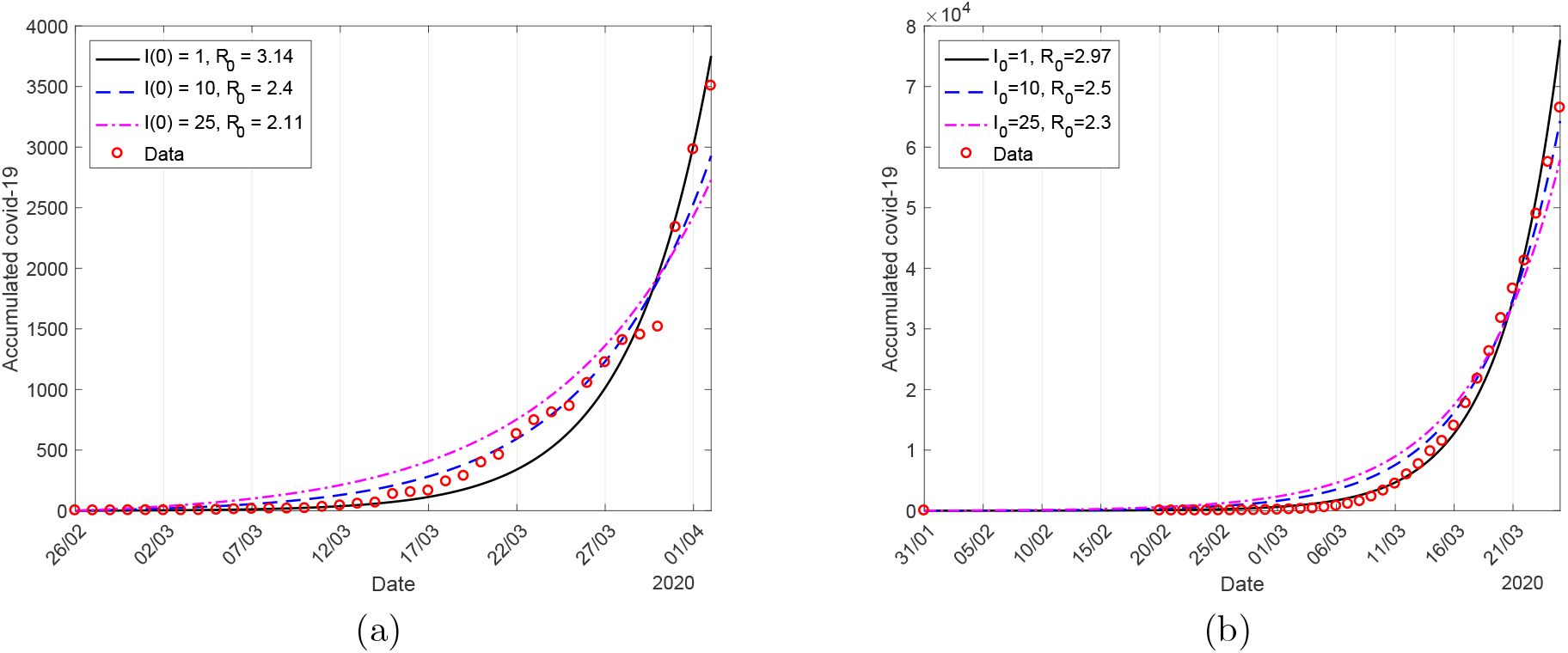
The estimated curve Ω for São Paulo (a) and Spain (b) with three different initial conditions *I*(0) = 1 (continuous curve), 10 (dashed curve), and 25 (dashed and dotted curve).

We observe that the larger the value of *I*(0), the small is the estimated *R*_0_. For instance, *R*_0_ with *I*(0) = 100 for São Paulo State is about half that with *I*(0) = 1. By the stringent definition of *R*_0_, we must consider *I*(0) = 1. However, the initial condition *I*(0) > 1 mimics the first case of CoViD-19 occurring earlier than the time *t* = 0. The Singapore University of Technology and Design [15] estimated *R*_0_ using *I*(0) = 100 for different countries, underestimating the basic reproduction number.

Let us illustrate the curves Ω, Ω_*d*_, *I* and *R*_*ef*_ for the SIR model in the natural (occurring without any control) epidemic. Equations (5), (14) and (15) are used to obtain Ω, Ω_*d*_ and *R*_*ef*_, and *I* is the solution of the system of equations (4). The inflection time calculated from 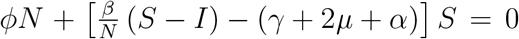 is *τ* = 78 (calendar time May 14). Figure 2 shows the estimated curve of Ω and the daily curve Ω_*d*_ (a), and the epidemic curve *I* and effective reproduction number *R*_*ef*_ (b). In Figure 2(a), Ω was divided by 10 to fit in the same frame with Ω_*d*_, and in 2(b), *I* was divided by 3 × 10^6^ to fit in the same frame with *R*_*ef*_. We used data collected from São Paulo State, and the initial conditions are *S*(0) = 44.6 million, *I*(0) = 1, and *R*(0) = 0, yielding *R*_0_ = 3.14.

**Figure 2:**
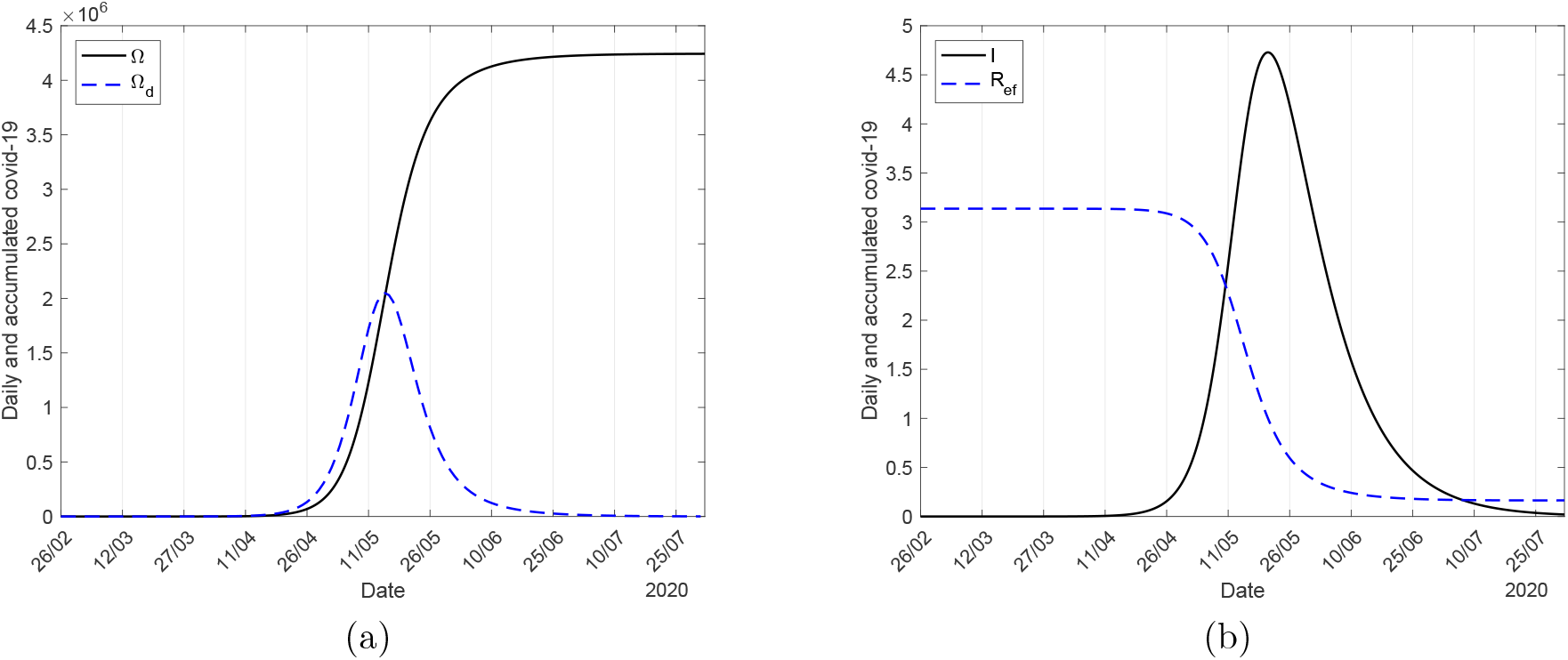
The estimated curve of Ω and the daily curve Ω_*d*_ (a), and the epidemic curve *I* and effective reproduction number *R*_*ef*_ (b).

The maximum value of Ω_*d*_ occurs at *t* = 78 (calendar time May 14), equal to the calculated inflection time *τ*. The maximum value of *I* occurs at *t* = 84 (calendar time May 20), equal to when *R*_*ef*_ = 1.

When *α* > 0, the steady-state fraction of the susceptible individuals *s*^*∗*^ is given by equation (A.10), and the effective reproduction number *R*_*ef*_ given by equation (15) is an approximated value. However, when *α* = 0, *s*^*∗*^ is given by equation (A.12), and *R*_*ef*_ = *R*_0_*S/N*, with *R*_0_ = *β/γ* + *ϕ* + *α*. Using the same model parameters values and initial conditions in Figure 2, the maximum values for Ω_*d*_ and *I* occurred at, respectively, *t* = 77 and 84, the inflection time is *τ* = 77, and *R*_*ef*_ = 1 occurred at *t* = 84. For *α* = 0, we have *R*_0_ = 3.20, which is 2% higher than *R*_0_ = 3.14. Therefore, *R*_*ef*_ = 1 occurs at the maximum value (peak) of the epidemic curve *I*.

### 3.2 The SEIR model

To estimate the transmission rate, we use equation (16) and Ω given by equation (3). The model parameters are those used in the SIR model: *γ* = 1*/*10, *α* = 0.002, and, for São Paulo State, *ϕ* = *µ* = 1*/*(78.4×365) and, for Spain, *ϕ* = *µ* = 1*/*(83.4×365) (all in *days*^−1^). For the additional parameter, we let *σ* = 1*/*5 *days*^−1^ (see Table 1). The transmission rate *β* is estimated, and the basic reproduction number *R*_0_ is calculated using equation (A.7). We estimate the basic reproduction number using equation (1) with the initial conditions *S*(0) = *N*_0_, *E*(0) = 0, *R*(0) = 0, and varying *I*(0).

For the data collected from São Paulo State, with *S*(0) = 44.6 million, we obtained for *I*(0) = 1, *R*_0_ = 7.25 with Sum = 1.36 × 10^6^, for *I*(0) = 10, *R*_0_ = 4.7 with Sum = 4.52 × 10^5^, for *I*(0) = 25, *R*_0_ = 3.82 with Sum = 1 × 10^6^, and for *I*(0) = 100, *R*_0_ = 2.6 with Sum = 3.68 × 10^6^. The lowest Sum occurs when *R*_0_ = 4.7.

For the data collected from Spain, with *S*(0) = 47.4 million, we obtained for *I*(0) = 1, *R*_0_ = 6.27 with Sum = 2.39×10^8^, for *I*(0) = 10, *R*_0_ = 4.75 with Sum = 7.64×10^7^, for *I*(0) = 25, *R*_0_ = 4.21 with Sum = 2.21 × 10^8^, and for *I*(0) = 100, *R*_0_ = 3.38 with Sum = 7.57 × 10^8^. The lowest Sum occurs when *R*_0_ = 4.75.

Figure 3 shows the estimated curve Ω for São Paulo (a) and Spain (b) with three different initial conditions (*I*(0) = 1, 10, and 25).

**Figure 3:**
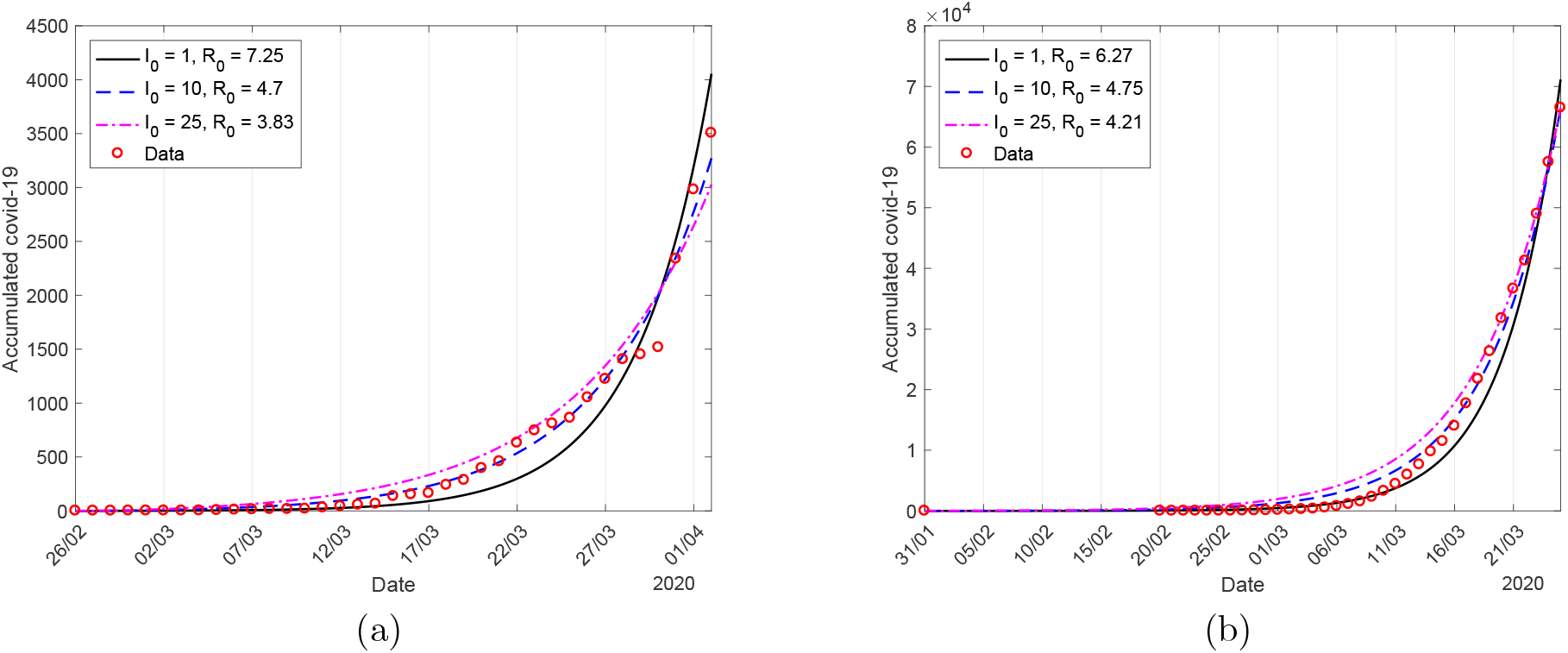
The estimated curve Ω for São Paulo (a) and Spain (b) with three different initial conditions *I*(0) = 1 (continuous curve), 10 (dashed curve), and 25 (dashed and dotted curve).

Notice that *σ/* (*σ* + *µ*) = 0.99999, hence, *R*_0_ given by equations (A.7) and (A.3) must be equal if *β* is the same for the SIR and SEIR models. However, *β* is estimated by the dynamic systems (4) or (1) against the accumulated severe CoViD-19 cases. For this reason, the estimated *β* must be different in the SIR and SEIR models.

Comparing with the estimated *R*_0_ provided by the SIR model, we observe that the SEIR model estimated with a higher value. Let us assess the role played by the incubation period *σ*^−1^ in the SEIR model considering the initial conditions *S*(0) = *N*_0_, *E*(0) = 0, *I*(0) = 1, and *R*(0) = 0. For *σ*^−1^ = 10 *days, R*_0_ = 11.27 with Sum = 1.15 × 10^6^, for *σ*^−1^ = 1 *day, R*_0_ = 3.92 with Sum = 8.92 × 10^5^, for *σ*^−1^ = 10^−1^ *days* (2.4 *hours*), *R*_0_ = 3.2 with Sum = 8.06 × 10^5^, and for *σ*^−1^ = 10^−3^ *days* (1.44 *minutes*), *R*_0_ = 3.14 with Sum = 7.96 × 10^5^. As *σ*^−1^ decreases (*σ* increases), *R*_0_ approaches (when *σ* → ∞) to that estimated by the SIR model (*R*_0_ = 3.14). By disregarding the incubation period, the SIR model provides a relatively lower estimation for *R*_0_ in comparison with the SEIR model. To overcome this period and fit the same set of the severe CoViD-19 cases, the transmission rate of SARS-CoV-2 must be much higher in the SEIR than in the SIR model. Therefore, the inclusion of the exposed individuals delays the onset of disease (or the entering into an infectious compartment), and the virus must infect more individuals (increased *R*_0_).

### 3.3 The SEAPMDR model

To estimate the transmission rates, we consider *β*_*y*_ = *β*_1*y*_ = *β*_2*y*_ = *β*_3*y*_ and *β*_*o*_ = *β*_1*o*_ = *β*_2*o*_ = *β*_3*o*_ = *ψβ*_*y*_, and we use equation (16) and Ω given by equation (11). The values for the model parameters are those given in Table 1. The basic reproduction number *R*_0_ is calculated using equation (12).

The initial conditions supplied to the system of equations (7), (8) and (9) are, for young and elder subpopulations,

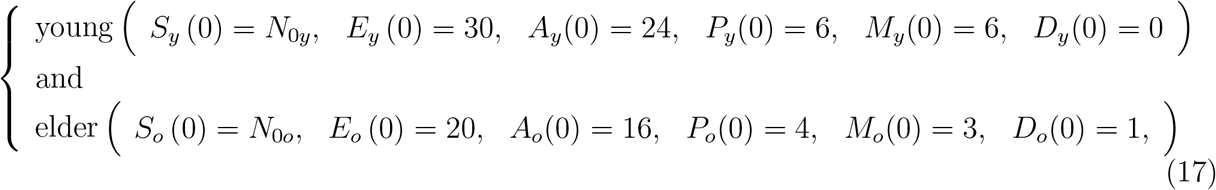

plus *R*(0) = 0, where the initial simulation time *t* = 0 corresponds to the calendar time when the first case was confirmed (February 26 for São Paulo State, and January 31 for Spain). For São Paulo State, *N*_0*y*_ = 37.8 million and *N*_0*o*_ = 6.8 million, and for Spain, *N*_0*y*_ = 35.17 million and *N*_0*o*_ = 12.23 million. (See [16] for details in the initial conditions’ setup.)

For the data collected from São Paulo State, we obtained *R*_0_ = 6.54, with Sum = 7.75 × 10^5^, while for the data collected from Spain, we obtained *R*_0_ = 5.88, with Sum = 1.1 × 10^8^. Figure 4 shows the estimated curve Ω for São Paulo State (a) and Spain (b).

**Figure 4:**
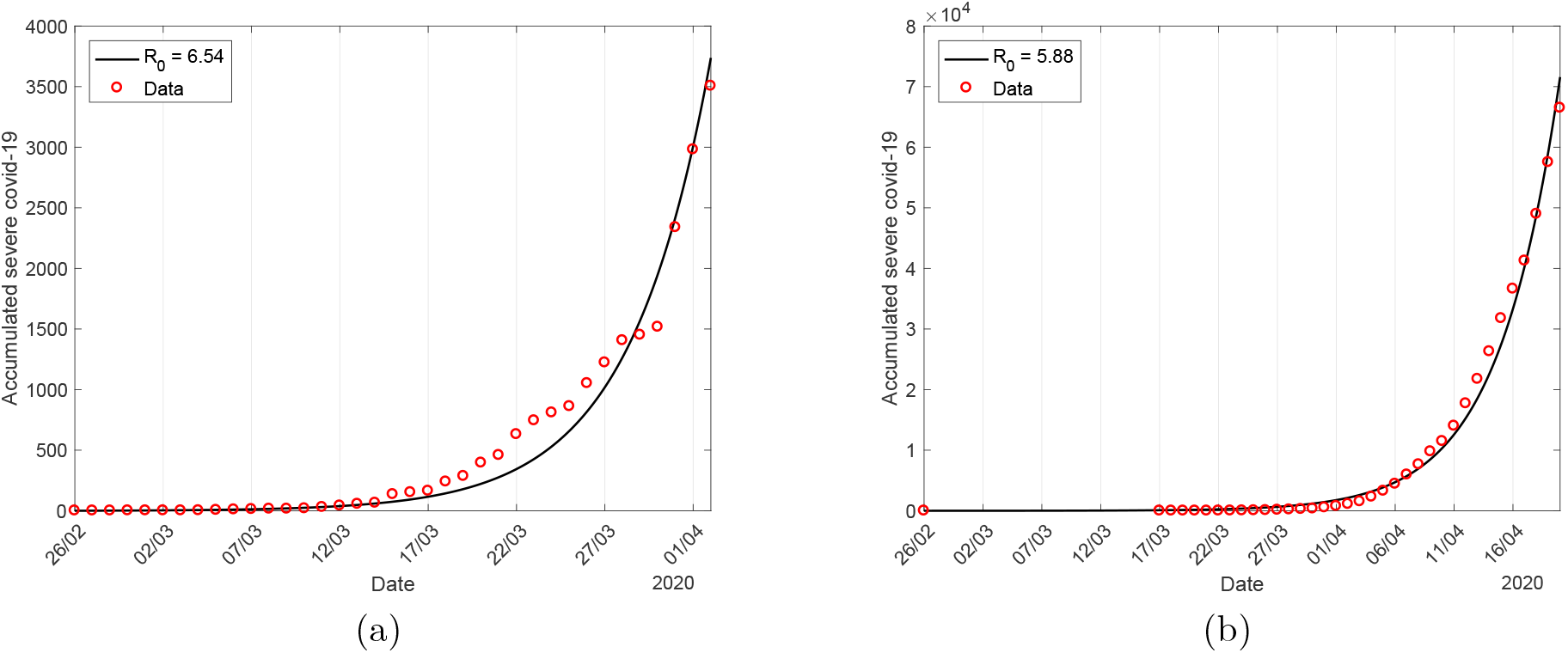
The estimated curve Ω and the observed accumulated cases for São Paulo State (a) and Spain (b).

Suppose we let *z*_*y*_ = *z*_*o*_ = 0 (mild CoViD-19 cases do not transmit) and *χ*_*y*_ = *χ*_*o*_ = 1 (asymptomatic individuals do not relapse to mild CoViD-19). In that case, the estimated basic reproduction number is *R*_0_ = 6.26 for São Paulo State, with Sum = 7.56 × 10^5^, and *R*_0_ = 5.67 for Spain, with Sum = 1.18 × 10^8^.

For the SEIR model, let us consider as the initial conditions *E*(0) = 50 (sum of *E*_*y*_ (0) + *E*_*o*_ (0)), *I*(0) = 1 and *R*(0) = 0, and *S*(0) = 44.6 million for São Paulo State and *S*(0) = 47.4 million for Spain. The estimated basic reproduction number for São Paulo State is *R*_0_ = 3.53 with Sum = 1.56 × 10^6^ and for Spain, *R*_0_ = 4.07 with Sum = 2.53 × 10^8^.

## 4 Discussion

There are different manners to define an epidemic curve. For instance, one possible definition is the curve formed by those positive for serological and PCR tests. However, in the early phase of the epidemic, the CoViD-19 epidemic curve must be defined by severe cases, which are the only available data. In the SEAPMDR model, the CoViD-19 epidemic curve was retrieved by estimating the transmission rates of asymptomatic, pre-diseased, and a fraction of mild classes. In [11], it was shown that the ratio between non-apparent (sum of compartments *E, A*, and *P*) and apparent (sum of compartments *M* and *D*) CoViD-19 is around 24, showing that SARS-CoV-2 is being transmitted by a huge number of hidden cases.

In the SEAPMDR model, the initial conditions *E*(0) = 50 and *I*(0) = 1 were supplied to the dynamic system, resulting in *R*_0_ = 6.54 (São Paulo State) and *R*_0_ = 5.88 (Spain), higher than estimations usually accepted. Instead of comparing the SIR model, let us compare the SEIR model with the same initial conditions supplied to the SEAPMDR model. The estimated basic reproduction number for São Paulo State was *R*_0_ = 3.92, and for Spain, *R*_0_ = 4.41.

Comparing SIR, SEIR, and SEAPMDR models, as the model incorporates more aspects of the natural history of the infection [17], higher becomes the estimation of *R*_0_. There is no other alternative for the SIR and SEIR models except considering severe CoViD-19 cases as infective. However, in the SEAPMDR model, the asymptomatic (*A*), pre-diseased (*P*), and a fraction of mild CoViD-19 (*M*) individuals are transmitting SARS-CoV-2, but the severe CoViD-19 (*D*) individuals are isolated and do not contribute, except to infect the hospital staff [11].

As we have pointed out, at the beginning and also in the early phase of the CoViD-19 epidemic, only hospitalized severe CoViD-19 cases were registered after the confirmation by serological and or PCR tests. These individuals are isolated in hospitals (receiving treatment) or discharged from hospitals but recommend being isolated in their homes. Then, somehow the majority of these individuals are not participating in the populational SARS-CoV-2 chain transmission.

In the SEAPMDR model, there are several infectious classes, but the severe CoViD-19 cases do not transmit the SARS-CoV-2, for this reason, *R*_0_ does not depend on the additional mortality rates *α*_*y*_ and *α*_*o*_ (see equations (12) and (13)). On the other hand, in the SIR and SEIR models, there is only one infectious class, and *R*_0_ depends on the additional mortality rate *α* (see equations (A.7) and (A.3)). Notice that in the SIR and SEIR models, the unique way to estimate the transmission rate is that severe covid-9 cases form the infective class *I*.

Besides the consideration of the CoViD-19 data, we discuss the magnitude of *R*_0_. Both SIR and SEIR models provided lower estimates for *R*_0_ than the SEAPMDR model.

As we have pointed out, Li *et al*. [3] estimated *R*_*ef*_ = 2.38 using the range of data recorded from 54 days after the epidemic’s onset to 16 days after the lockdown. From Figure 2b corresponding to the SIR model with *R*_0_ = 3.14 (simulation time *t* = 0), we observe that *R*_*ef*_ *R*_0_ up to the simulation time *t* = 50 initiates a quick decreasing phase at *t* = 60. However, for the SEAPMDR model without interventions, *R*_0_ = 6.54 decreases to *R*_*ef*_ = 5.41 at simulation time *t* = 50. However, the partial quarantine was introduced on March 24 (*t* = 27), and 16 days later on April 9 (*t* = 45), we have *R*_*ef*_ = 2.92 considering protective measures in the SEAPMDR model (see [16]). This value is close to that estimated by Li *et al*., remembering that they did not take the initial time of estimation when the first case of CoViD-19 occurred.

The curve of accumulated CoViD-19 cases Ω obtained from equation (11) shown in Figure 2 has a sigmoid-shape. This curve presents a quick increase during the first phase (*R*_*ef*_ > 1, with upward concavity) followed by a slow increase (*R*_*ef*_ < 1, with downward concavity), where *R*_*ef*_ is given by equation (15). Figure 2 showed Ω and *R*_*ef*_ corresponding to the natural epidemic. In Figure 5, we show the daily Ω_*d*_ and the accumulated severe CoViD-19 cases Ω^*ob*^ collected from São Paulo State (a) and Spain (b), where *A* indicates the time at which quarantine was introduced, and *B* represents the inflection time. To be fitted together in the same frame, the accumulated data set was divided by 4. The observed severe CoViD-19 data carry on the effects of the quarantine and the protective measures (face mask, washing hands with alcohol and gel, social distancing, etc.) adopted by individuals that reduced the transmission of SARS-CoV-2. In São Paulo State, a partial quarantine was introduced on March 24 isolating approximately 53% of the population [8], and a rigid lockdown in Spain was introduced on March 16 isolating perhaps 90% or more.

**Figure 5:**
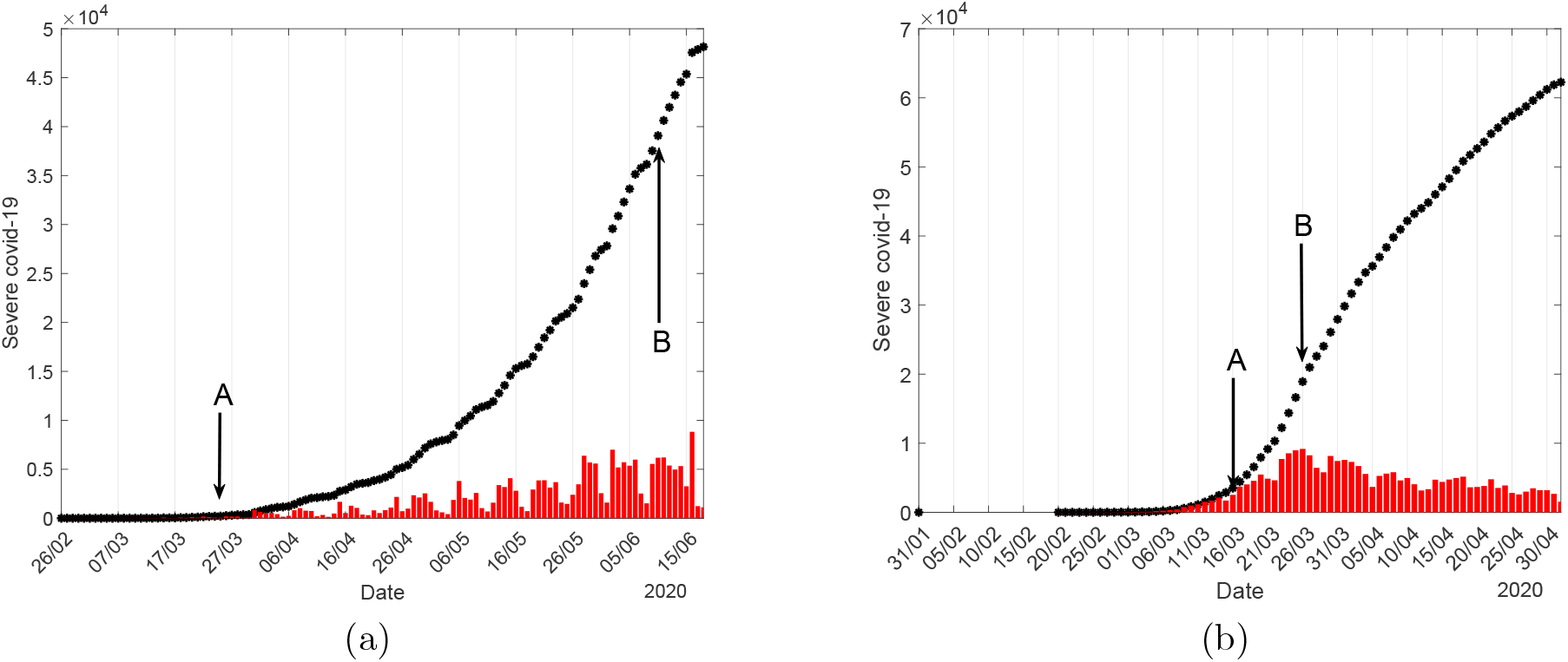
The daily (bars) and the accumulated (points) severe covid-19 cases in São Paulo State (a) and Spain (b), where *A* indicates the time at which quarantine was introduced, and *B* indicates the inflection time.

Let us define the threshold of the proportion in isolation in a population as *q*^*th*^ = 1 − 1*/R*_0_. If the proportion in quarantine *q* is higher than *q*^*th*^, we must have *R*_*ef*_ < 1, and the observed accumulated CoViD-19 cases must present downward concavity and upward concavity if *q* < *q*^*th*^. For São Paulo State, we have *q* = 0.53. At *R*_*ef*_ = 1, we have the inflection time (change from upward to downward concavity). Using the value of *R*_0_ estimated in the preceding section, for the SIR model (*R*_0_ estimated with *I*(0) = 1) we have *q*^*th*^ = 0.68 for São Paulo State, and *q*^*th*^ = 0.66 for Spain. For the SEAPMDR model, we have *q*^*th*^ = 0.85 for São Paulo State, and *q*^*th*^ = 0.83 for Spain. The SEAPMDR model shows that isolating 85% in the population or more controls the epidemic, but the SIR model predicts less proportion of isolation, 68% or more.

As shown in Figure 5, the inflection time occurred approximately on June 10 in São Paulo State and on March 26 in Spain. The elapsed time between the implementation of quarantine (*A*) and the inflection time (*B*) is 78 *days* in São Paulo State and 10 *days* in Spain. From Figure 2, for São Paulo State using the SIR model, the period from the beginning of the epidemic until the inflection time was 78 *days*. This period, which was calculated for the natural epidemic, must be lesser when considering the epidemic under isolation and protective measures. Hence, the actual *R*_0_ must be higher than 3.14. However, if we consider *R*_0_ = 2.11 estimated from the SIR model using *I*(0) = 25, we obtain *q*^*th*^ = 0.53 for São Paulo State resulting in *q* = *q*^*th*^. In other words, the inflection time must occur 9 days later, when we have *R*_*ef*_ = 1 (the effects of any intervention appear 9 days later). As a consequence, the observed accumulated severe CoViD-19 cases must be quite similar to that observed in Spain. Notice that the inflection time occurred 10 days after the implementation of lockdown in Spain. The long time to reach the inflection time in São Paulo State (78 days) may indicate that the SIR model underestimated *R*_0_, and that estimated by the SEAPMDR model seems to be more reliable.

Finally, the SEAPMDR model proposed by Yang *et al*. [6] at the beginning of the CoViD-19 epidemic deserves some considerations. At the beginning of the epidemic, the severe acute respiratory syndrome cases were hospitalized and confirmed as CoViD-19 after some tests (many patients died and were diagnosed *post mortem* as CoViD-19 by a test). This disease’s rapid spread indicated that possibly asymptomatic and mild CoViD-19 cases transmitted SARS-CoV-2 once severe CoViD-19 patients were isolated in hospitals. Additionally, the model considered pre-symptomatic individuals transmitting infection, which was confirmed later. Besides these aspects, the model considered the increased fatality among elder individuals. Minimalist incorporation of this disease’s severity in the model divided the population into two groups: young (under 60 years old) and elder (above 60 years old) individuals. (Notably, the consideration of two subpopulations by the SEAPMDR model predicted three times lower number of deaths than that indicated in [2].) To estimate the basic reproduction number, the authors restricted data collection during the period without any control.

## 5 Conclusion

This work’s goal was to demonstrate that the early applied SIR and SEIR models underestimated *R*_0_ misusing the severe CoVid-19 data collection. For this reason, we formulated the SEAPMDR model incorporating essential aspects related to the natural history of the infection. For instance, the incorporation of the asymptomatic and pre-diseased individuals, mild CoViD-19 cases, and different fatality rates depending on age must improve the mathematical model to describe the CoViD-19 epidemic. Additionally, these more elaborated models could consider the severe CoViD-19 cases being isolated, and SARS-CoV-2 is transmitted by asymptomatic and pre-diseased individuals, for instance. Hence, the SEAPMDR must provide a more accurate estimation of *R*_0_. Specifically, when the severe CoViD-19 cases may not transmit SARS-CoV-2 populationally, the SIR and SEIR models structured in only one infectious compartment are not suitable to estimate the basic reproduction number *R*_0_. It is worth stressing that the reliable estimation of *R*_0_ must consider the time range from the first case to the last case before the first case affected by interventions.

## Data Availability

Data obtained from ministery of health fo Spain and São Paulo State.

https://www.seade.gov.br/coronavirus/

https://www.mscbs.gob.es/profesionales/saludPublica/ccayes/alertasActual/nCov/home.htm

## Availability of data and material

The data that support the findings of this study are openly available in SP contra o novo coronavírus (Boletim completo) at https://www.seade.gov.br/coronavirus/, and Spain Covid19 at https://cnecovid.isciii.es/covid19/.

## Author contributions

**Hyun Mo Yang**: Conceptualization, Methodology, Formal analysis, Writing - Original draft preparation, Validation, Supervision. **Luis Pedro Lombardi Junior**: Software, Data Curation, Visualization, Validation. **Ariana Campos Yang**: Conceptualization, Validation, Investigation.

## A The steady-state analysis of the SIR and SEIR models

We present the analysis of the SIR and SEIR models.

### A.1 The SEIR model

The system of equations (1) does not reach a steady-state, except if *ϕ* = *µ* + *αI/N*. However, the system (1) in terms of the fractions attains steady-state. Defining the fraction *x* = *X/N*, with *X* = {*S, E, I, R*}, we have

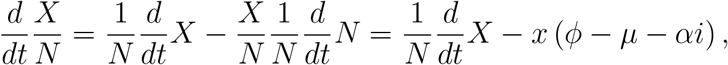

using equation (2), and the system of equations (1) becomes

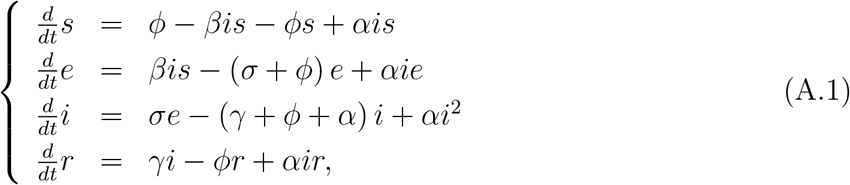

with *s* + *e* + *i* + *r* = 1, hence, the equation for *r* can be decoupled from the system, through *r* = 1 − *e* − *s* − *i*. Notice that *d* (*s* + *e* + *i* + *r*) */dt* = 0, and the system of equations in terms of fractions attain a steady state.

The system of equations (A.1), dropping out the decoupled equation for *r*, has two equilibrium points: The trivial (disease-free) equilibrium point 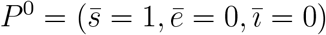 and non-trivial (epidemic) equilibrium point 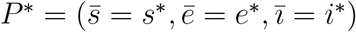.

Let us assess the stability of *P* ^0^ by applying the next generation matrix theory considering the vector of variables *x* = (*e, i*) [18]. The next generation matrix is constructed considering a subsystem of equation (9) taking into account the state-at-infection (*e*) and the states-of-infectiousness (*i*) [18], resulting in *x* = (*e, i*). In a matrix form, the subsystem is written as

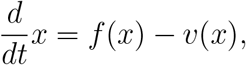

where the vectors *f* and *v* are defined below, with the partial derivatives of *f* and *v* evaluated at *P*^0^ being given by

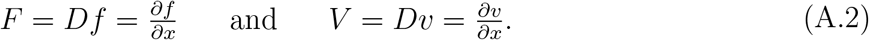

Depending on the choice of vectors *f* and *v*, we can obtain the reduced reproduction number or the fraction of susceptible persons at endemic level [19].

To obtain the basic reproduction number *R*_0_, diagonal matrix *V* is considered. Hence, the vectors *f* and *v* are

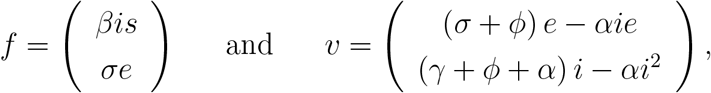

resulting in *F* and *V* given by

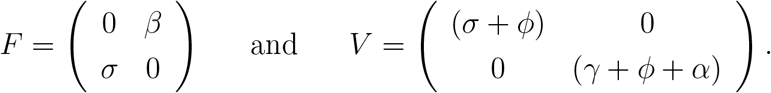

The characteristic equation corresponding to *FV* ^−1^ is

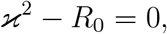

and the spectral radius 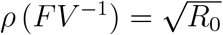, where the basic reproduction number *R*_0_ is

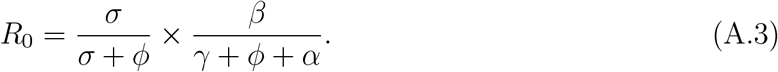

Instead of the spectral radius 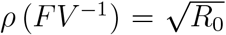, we applied the procedure in [19] (the sum of coefficients of the characteristic equation) and proved in [20], resulting in the threshold *R*_0_. Hence, the trivial equilibrium point *P* ^0^ is locally asymptotically stable (LAS) if *R*_0_ < 1.

To obtain the fraction of susceptible individuals at endemic equilibrium *s*^*∗*^, *F* must be the most straightforward (matrix with the least number of non-zero elements). Hence, the vectors *f* and *v* are

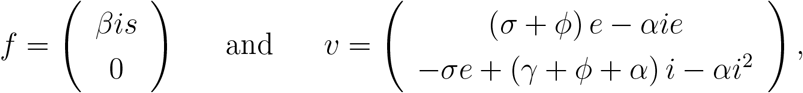

resulting in *F* and *V* given by

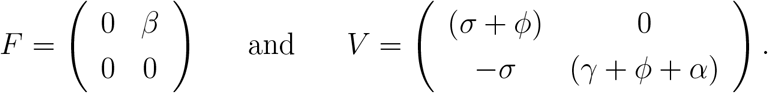

The characteristic equation corresponding to *FV* ^−1^ is

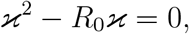

and the spectral radius *ρ* (*FV* ^−1^) = *R*_0_, which is equal to the sum of the coefficients.

Both procedures resulted in the same threshold, hence, according to [21], the inverse of the reduced reproduction number *R*_0_ given by equation (A.3) is a function of the fraction of susceptible individuals at endemic equilibrium *s*^*∗*^ through

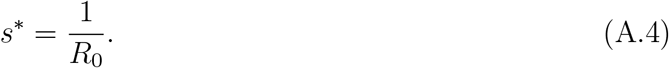

Hence, we can define the effective reproduction number *R*_*ef*_ as

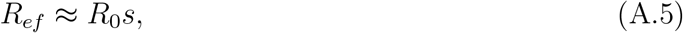

which depends on time, with *R*_*ef*_ = *R*_0_ at *t* = 0 (*s* = 1), and when attains steady-state (*R*_*ef*_ = 1), we have *s*^*∗*^ = 1*/R*_0_. Notice that *R*_*ef*_ = *R*_0_*s* when *α* = 0 (see below for the SIR

model),

### A.2 The SIR model

The SIR model is obtained letting *σ* → ∞ in the SEIR model, and the system of equations (4) becomes

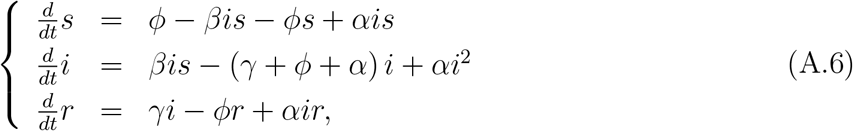

with *s* + *i* + *r* = 1, hence, the equation for *r* can be decoupled from the system, through *r* = 1 − *s* − *i*. The system of equations (A.6), dropping out the decoupled equation for *r*, has two equilibrium points: The trivial (disease-free) equilibrium point 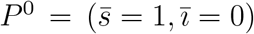 and non-trivial (epidemic) equilibrium point 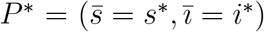. The stability analysis of trivial equilibrium is given from the preceding section letting *σ* → ∞, and the the basic reproduction number *R*_0_ is

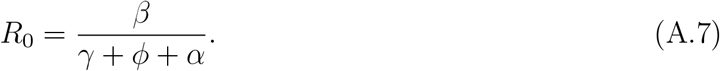

The coordinates of the non-trivial equilibrium point 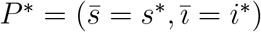 in the SIR model can be calculated by

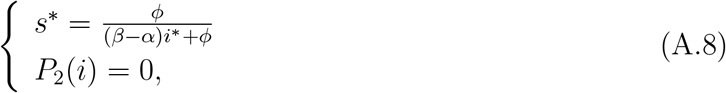

where *i*^*∗*^ is the positive root but small than one of the second degree polynomial *P*_2_(*i*) given by

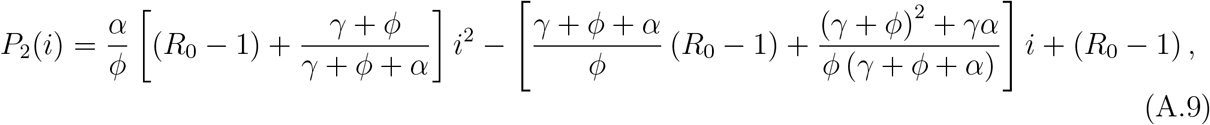

which has the value, at *i* = 1,

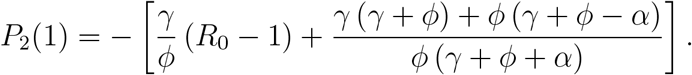

When *R*_0_ > 1, we have *P*_2_(1) < 0 (the condition *γ* + *ϕ* > *α* is satisfied because *γ* > *α*), and the two positive roots of *P*_2_(*i*) are such that 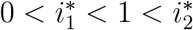. Hence the small root 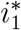 is biologically feasible. When *R*_0_ = 1, we have 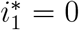 and 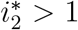, hence 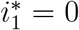 is biologically feasible. When *R*_0_ < 1, we have 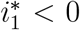 and 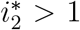, hence 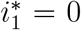 is biologically feasible. Therefore, the small root 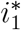, which is biologically feasible, assumes a negative value for *R*_0_ < 1, zero at *R*_0_ = 1, and a positive value but lower than 1 for *R*_0_ > 1. The small root of *P*_2_(*i*) is given by

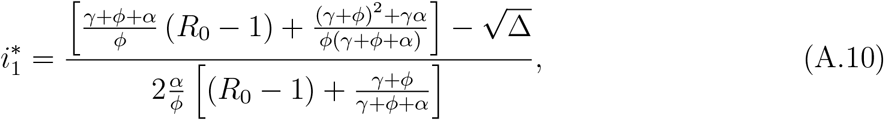

where Δ is

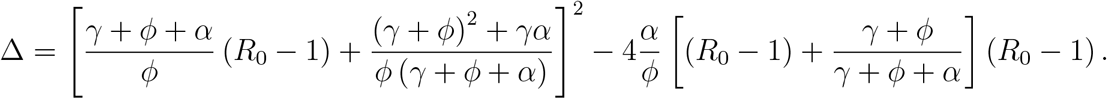

The complexity arises due to the non-constant population under the additional mortality rate. Let us consider *α* = 0. In this case, *P*_2_(*i*) has a unique positive solution

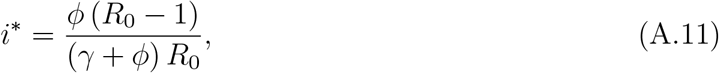

and the fraction of susceptible individuals, from equation (A.8), is

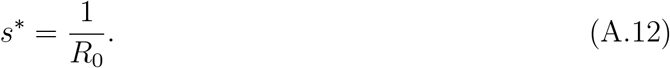

In this particular case, we have *R*_*e*_ = *R*_0_*s*. Hence, for *α* > 0, comparing equations (A.8) and (A.10), we notice that *s*^*∗*^ has a complex dependency with *R*_0_, not simply 1*/R*_0_.

Let us assess the stability of the system of equations (A.6) linearized around the trivial equilibrium point 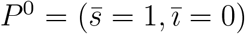. The Jacobian matrix *J* evaluated at the trivial equilibrium point *P* ^0^ is

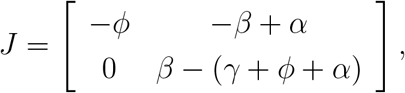

with the eigenvalues *ρ*_1_ = −*ϕ* and *ρ*_2_ = (*γ* + *ϕ* + *α*) (*R*_0_ − 1), where the basic reproduction number *R*_0_ is given by (A.7). Hence, *P* ^0^ is locally asymptotically stable if *R*_0_ < 1. Hence, the linearized system around *P* ^0^ has a trajectory for *i* given by

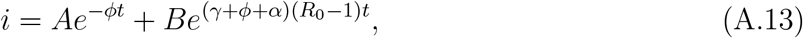

where *A* and *B* are arbitrary values determined by the initial conditions.

When *R*_0_ > 1, an introduction of one infectious individual disestablishes the trivial equilibrium point *P* ^0^, and the unique positive eigenvalue gives the leaving trajectory. Hence, the escaping trajectory follows 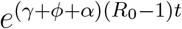in the vicinity of *P* ^0^ towards the non-trivial equilibrium point *P*^*∗*^. Maidana and Yang [22] provided an example of trajectories (travelling waves) linking *P* ^0^ towards *P*^*∗*^ considering a spatial model for the dengue transmission.

To establish a relationship with the linearized system of equations in terms of the fractions, the system of equations (4), using *R* = *N* − *I* − *S*, is rewritten as

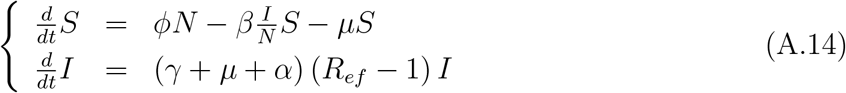

where the effective reproduction number *R*_*ef*_ is defined by

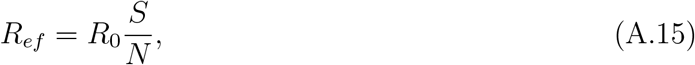

with the basic reproduction number *R*_0_ being given by equation (A.7) changing *ϕ* by *µ*, that is, *R*_0_ = *β/* (*γ* + *µ* + *α*).

Let us analyze the system of equations (A.14) at two boundaries. Let us assume that the first case of CoViD-19 is introduced at *t* = 0, that is, the initial conditions supplied to equation (A.14) are *S*(0) = *N* − 1 and *I*(0) = 1. For a large population, we can approximate *S* ∼ *N*, and the system of equations can be approximated by

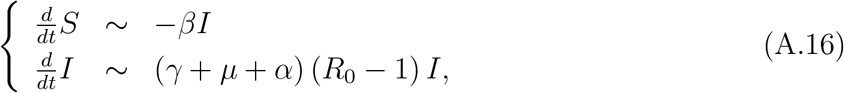

with *R* = *N* − *S* − *I*, and at the beginning of the epidemic, if we estimate the transmission rate *β*, we can calculate *R*_0_ using the expression obtained from the steady-state analysis. The system of equations (A.14) does not approach to a steady-state, but attains it when *α* = 0. In this case, asymptotically (*t* → ∞), we have *dI/dt* = 0 if *R*_*ef*_ = *R*_0_*S/N* = 1, that is, *S* → *S*^*∗*^ = *s*^*∗*^*N* and *I* → *I*^*∗*^ = *i*^*∗*^*N*, where *i*^*∗*^ and *s*^*∗*^ are given by equations (A.11) and (A.12), respectively. Hence, when *α* = 0, at *t* = 0, *R*_*ef*_ = *R*_0_, and when *t* → ∞ (steady-state), *R*_*ef*_ = 1 (see equation (A.12)), from which we retrieve the well known relationship *s*^*∗*^ = 1*/R*_0_ [4].

Therefore, based on *R*_*ef*_ given by equation (A.15) when *ϕ* = *µ* and *α* = 0, the basic reproduction number *R*_0_ obtained from mathematical modelings provides two useful information: At the beginning of the epidemic (*t* = 0), *R*_0_ gives the magnitude of the initial takeoff of the epidemic, and when epidemic reaches the steady-state (after many waves of the epidemic, that is, *t* → ∞), *R*_0_ measures its severity providing the fraction of susceptible individuals, that is, *s*^*∗*^ = 1*/R*_0_. Between these two extremes, the effective reproduction number *R*_*ef*_ dictates the course of an epidemic, which follows decaying oscillations around *R*_*ef*_ = 1 [7]. It is worth stressing that *R*_*ef*_ given by equation (A.15) is valid only when *ϕ* = *µ* and *α* = 0, and when one of these conditions is not valid, *R*_*ef*_ given by equation (A.15) can be used as an approximated value.

From equation (A.16), the solution for *I* is 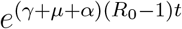, which is equal to equation (A.13) when *A* = 0 and *B* = 1.

## B The steady-state analysis of the SEAPMDR model

The system of equations (7), (8) and (9) does not reach steady state, except if *ϕ* = *µ* + (*α*_*y*_*D*_*y*_ + *α*_*o*_*D*_*o*_) */N*, when the total size of the population is constant. However, the system of equations (7), (8) and (9) in term of fractions attains steady-state. Defining the fraction *x*_*j*_ = *X*_*j*_*/N*, for *j* = *y, o*, with *X*_*j*_ = {*S*_*j*_, *E*_*j*_, *A*_*j*_, *P*_*j*_, *M*_*j*_, *D*_*j*_, *R*}, we have

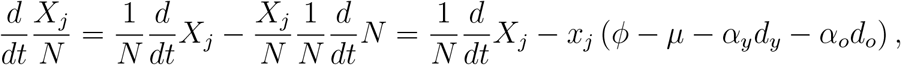

using equation (10), and the system of equations (7), (8) and (9) in terms of fractions become, for susceptible individuals,

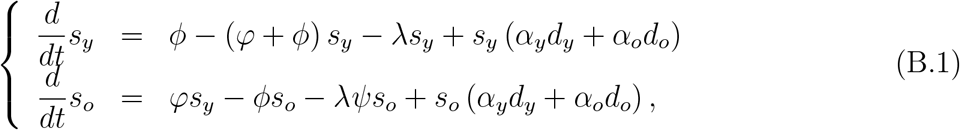

for infected individuals,

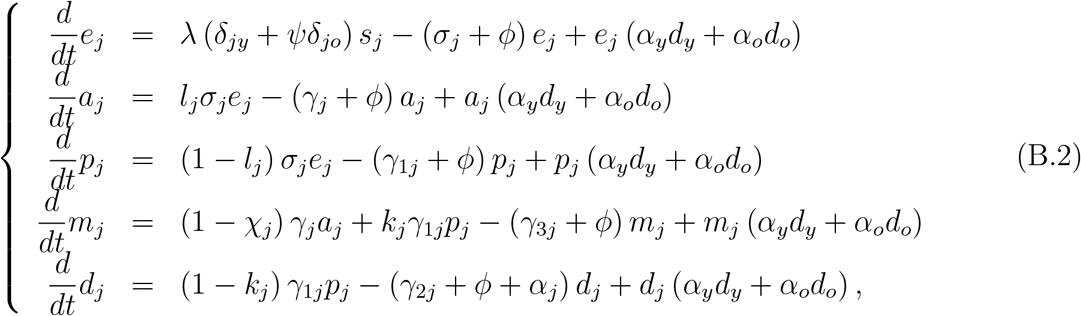

and for recovered individuals,

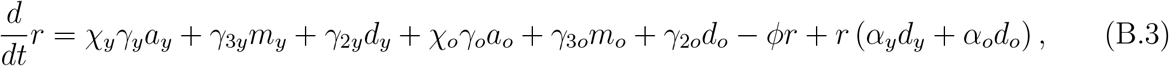

where *λ* is the force of infection given by equation (6) re-written as

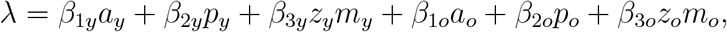

and

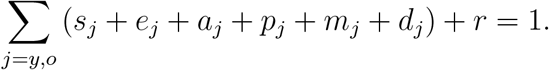

This new system of equations has a steady-state, that is, the number of individuals in all classes varies with time. However, their fractions attain a steady-state (the sum of derivatives of all classes is zero).

The trivial (disease-free) equilibrium point *P* ^0^ of the new system of equations (B.1), (B.2) and (B.3) is given by

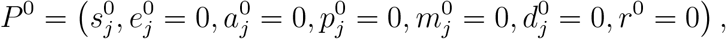

for *j* = *y* and *o*, where

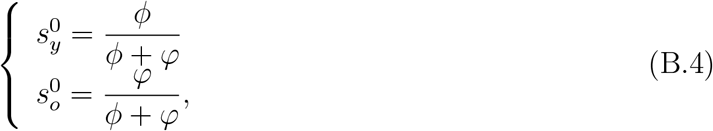

with 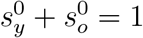.

Let us assess the stability of *P* ^0^ by applying the next generation matrix theory considering the vector of variables *x* = (*e*_*y*_, *a*_*y*_, *p*_*y*_, *m*_*y*_, *e*_*o*_, *a*_*o*_, *p*_*o*_, *m*_*o*_) [18]. We apply method proposed in [19] and proved in [20]. To obtain the basic reproduction number, diagonal matrix *V* is considered. Hence, the vectors *f* and *v* are

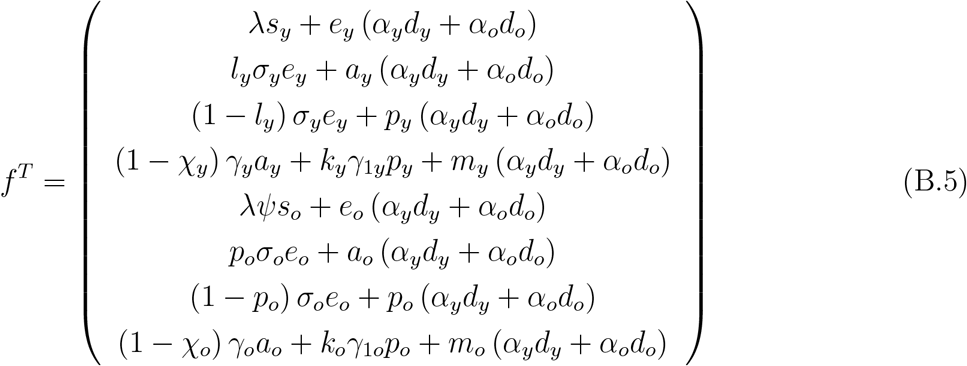

and

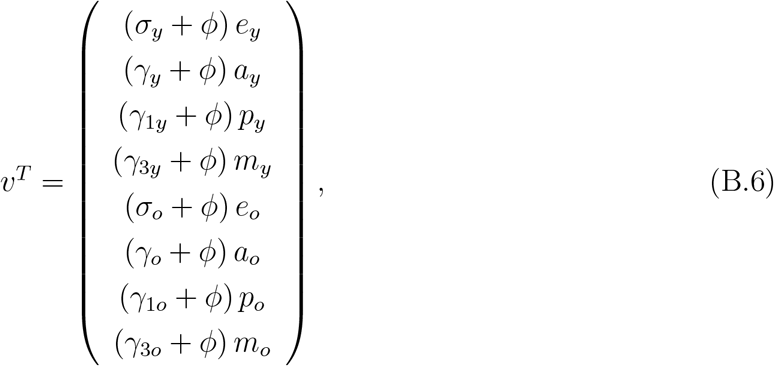

where the superscript *T* stands for the transposition of a matrix, from which we obtain the matrices *F* and *V* (see [18]) evaluated at the trivial equilibrium *P* ^0^, which were omitted. The next generation matrix *FV* ^−1^ is

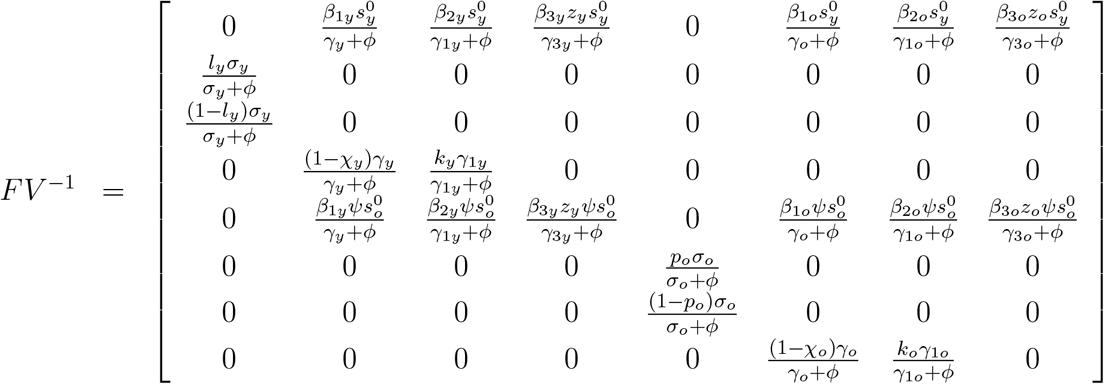

and the characteristic equation corresponding to *FV* ^−1^ is

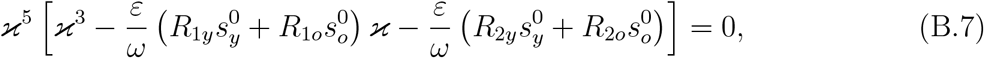

with the basic reproduction number *R*_0_ being given by

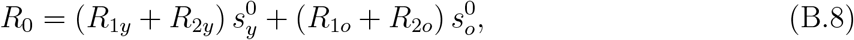

where the initial fractions 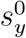 and 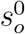 are given by equation (B.4), and the partial basic reproduction numbers *R*_1*y*_, *R*_2*y*_, *R*_1*o*_, and *R*_2*o*_ are given by equation (13) in the main text. The spectral radius *ρ* (*FV* ^−1^) is the biggest solution of a third-degree polynomial, not easy to evaluate. The procedure proposed in [19] allows us to obtain the threshold *R*_0_ as the sum of coefficients of the characteristic equation, where *R*_0_ is the basic reproduction number given by equation (12) in the main text. Hence, the trivial equilibrium point *P* ^0^ is locally asymptotically stable if *R*_0_ < 1.

